# Selecting COVID-19 Convalescent Plasma for Neutralizing Antibody Potency Using a High-capacity SARS-CoV-2 Antibody Assay

**DOI:** 10.1101/2020.08.31.20184895

**Authors:** E Goodhue Meyer, G Simmons, E Grebe, M Gannett, S Franz, O Darst, C Di Germanio, M Stone, P Contestable, A Prichard, R Reik, R Vassallo, P Young, MP Busch, P Williamson, LJ Dumont

## Abstract

**BACKGROUND:** Efficacy of COVID-19 convalescent plasma (CCP) to treat COVID-19 is hypothesized to be associated with the concentration of neutralizing antibodies (nAb) to SARS-CoV-2. High capacity serologic assays detecting binding antibodies (bAb) have been developed, nAb assays are not adaptable to high-throughput testing. We sought to determine the effectiveness of using surrogate bAb signal-to-cutoff ratios (S/CO) in predicting nAb titers using a pseudovirus reporter viral particle neutralization (RVPN) assay.

**METHODS:** CCP donor serum collected by 3 US blood collectors was tested with a bAb assay (Ortho Clinical Diagnostics VITROS Anti-SARS-CoV-2 Total, CoV2T) and a nAb RVPN assay. CoV2T prediction effectiveness at S/CO thresholds was evaluated for RVPN nAb NT_50_ titers using receiver operating characteristic analysis.

**RESULTS:** 753 CCPs were tested with median CoV2T S/CO of 71.2 and median NT_50_ of 527.5. Proportions of CCP donors with NT_50_ over various target nAb titers were 86% ≥1:80, 76% ≥1:160, and 62%≥1:320. Increasing CoV2Ts reduced the sensitivity to predict NT_50_ titers, while specificity to identify those below thresholds increased. As the targeted NT_50_ increased, the positive predictive value fell with reciprocal increase in negative predictive value. S/CO thresholds were thus less able to predict target NT_50_ titers.

**CONCLUSION:** Selection of a clinically effective nAb titer will impact availability of CCP. Product release with CoV2T assay S/CO thresholds must balance the risk of releasing products below target nAb titers with the cost of false negatives. A two-step testing scheme may be optimal, with nAb testing on CoV2T samples with S/COs below thresholds.

## Introduction

A novel coronavirus, SARS-CoV-2, first identified as the causative agent of a late 2019 outbreak of atypical pneumonia in Wuhan, China, was declared a pandemic by the World Health Organization in March 2020. It is not only the high transmissibility of SARS-CoV-2 that made it a significant public health concern, but also the heterogeneity and seriousness of clinical manifestations in the form of COVID-19 disease. There had been over 20 million confirmed cases worldwide and more than 5 million in the United States by August 25^th^. The death toll worldwide had reached in excess of 814,000, with the United States accounting for over 20% of those deaths (~177,773).^1^

Over the past 6 months the medical and scientific community has worked to characterize the pathologic course and determine the best treatment options for COVID-19. Currently viral specific therapies and vaccines are under development. The most specific therapeutic option currently available is an old standby: convalescent plasma (CP). On March 24, 2020, the Food and Drug Administration released a pathway for investigational new drug (IND) use of COVID-19 convalescent plasma (CCP) for the treatment of patients suffering from severe, life-threatening COVID-19.^2^ Clinical use of CCP has expanded rapidly through the Mayo Clinic’s Expanded Access Program (EAP), single-patient emergency INDs, and single or multi-center institutional INDs.^2^ Several observational studies have demonstrated clinical benefit from 1-2 unit transfusions of CCP.^3–5^ An early report for the Mayo Clinic EAP showed reduced mortality in patients who received CCP early (within 3 days of diagnosis) and a mortality benefit with higher IgG antibody levels in the transfused plasma.^6^ The safety of CCP transfusions was similar to standard plasma transfusions.^7^ A randomized controlled trial conducted in Wuhan, however, did not report improved outcomes with CCP treatment for patients with severe COVID-19, although the study was terminated early due to a decrease in the COVID-19 case load.^8^

The transfusion community mobilized efforts to recruit and collect CCP from recovered patients in the context of rapidly evolving FDA guidance and testing technologies. In previous outbreaks, the clinical benefit of CP was related to the dose of neutralizing antibodies in the plasma.^9,10^ Neutralizing antibodies directly block viral attachment to the host cell and subsequent fusion.^11,12^ A small 2018 study on the use of CP in a Middle East Respiratory Syndrome outbreak demonstrated that the neutralizing titer of the convalescent plasma should exceed 1:80 to be clinically effective.^10^ The CCP used in Duan et al.’s retrospective review of 10 patients with severe COVID-19 and Shen et al.’s of 5 patients had high titers (ie. >1:640).^3,4^ Despite some encouraging data, the clinical utility of CCP in COVID-19 has not been established, and the optimal dose and types of anti-SARS-CoV-2 antibodies in CCP remain uncertain.

Current FDA guidance through the CCP Emergency Use Agreement (EUA) guidelines recommends high titer CCP with an S/CO of >12 with the Ortho VITROS Anti-SARS-CoV-2 Ig only assay (Ortho Clinical Diagnostics, Raritan, New Jersey) which correlates to a neutralizing nAb titer of >1:250.^13^ FDA has recognized that qualification of CCP products on the basis of nAb titer is challenging, given that conventional neutralizing antibody titer assays are cell-based, not amendable to high throughput testing, and not standardized. To that end on April 20, 2020, Creative Testing Solutions (CTS) implemented the VITROS Anti-SARS-CoV-2 Total Ig assay (CoV2T, Ortho Clinical Diagnostics, Raritan, New Jersey), which had received FDA emergency use authorization (EUA) on April 3, 2020. The assay detects binding antibodies (bAb) to the S1 subunit of the SARS-CoV-2 spike glycoprotein.

Three large blood collection organizations began using the CoV2T semiquantitative test to ascertain donor seroreactivity to qualify CCP products, although the correlation of CoV2T signal-to-cutoff ratio (S/CO) with SARS-CoV-2 nAb titers was unknown. In this study we endeavored to determine the effectiveness of CoV2T S/CO in predicting titers of nAbs to the immunodominant S protein, measured using SARS-CoV-2 Reporter Viral Particle Neutralization (RVPN) assay (Vitalant Research Institute (VRI), San Francisco, CA). We evaluated the reliability of a range of CoV2T S/CO thresholds for predicting nAb titer, in an effort to derive optimal testing algorithms for CCP qualification.

## Materials and Methods

We tested serum samples from 753 CCP donations collected by the American Red Cross (ARC), OneBlood and Vitalant during April and May 2020 with both the Ortho VITROS® CoV2T serologic assay and the VRI RVPN assay.

### CCP Donor Qualification

CCP donor collection began in early April 2020. ARC study donors were collected from 4/8/20-5/19/20, OneBlood from 3/30/2020 – 5/12/2020, and Vitalant from 4/12/2020-5/13/220. FDA mandated CCP donor qualification evolved throughout the study period due to testing availability and evolution of the pandemic in the United States. Evidence of COVID-19 was required in the form of a documented positive SARS-CoV-2 molecular or serologic test, and either complete resolution of symptoms at least 28 days prior to donation or complete resolution 14 days prior to donation with negative molecular test result. All CCP donors were also required to meet traditional allogeneic blood donor criteria per the Code of Federal Regulations (21 CFR 630.10 and 630.15).^14^ At time of plasma collection, donors consented to use of de-identified donor information and test results for research purposes. ARC collected apheresis plasma units on Alyx and Amicus devices (Fresenius Kabi USA LLC, Lake Zurich, IL). At OneBlood, whole blood plasma collection was performed using the Haemonetics Leukotrap blood collection system (Haemonetics, Braintree, Mass.). Apheresis plasma collection was performed at both OneBlood and Vitalant using Alyx devices and Trima (Terumo BCT, Lakewood, CO) apheresis systems following manufacturers’ recommendations.

On April 27, 2020, ARC began CCP donor qualification with the CoV2T assay. Units from donors with a signal to cutoff (S/CO) ratio ≥1.0 (reactive per package insert) were labeled as CCP units. Retrospective testing was accomplished for the donors collected from 4/8/2020 – 4/26/2020 from frozen serum drawn at time of collection. Vitalant began CoV2T testing on 5/6/2020 but start-up reporting delays precluded its use as a release criterion until after 5/13/2020 with retrospective testing on previously frozen samples completed thereafter. Vitalant also used an S/CO ≥1.0 as a CCP unit release criterion. Vitalant has allowed repeat CCP donations every 7 days.

### Preparation of Samples

Either refrigerated serum from the routine donation testing tubes, or frozen serums from randomly selected CCP donations were sent to the CTS in St. Petersburg, Florida. Serum aliquots were prepared at CTS for CoV2T testing by individual donation. Regardless of VITROS Anti-SARS-CoV-2 Total Ab reactivity status, the serums aliquots were refrozen after testing and then shipped on dry ice to Vitalant Research Institute (VRI; San Francisco, CA) for assessment of the sample's SARS-CoV-2 neutralization activity.

### Serologic Testing

VITROS Anti-SARS-CoV-2 Total testing was performed following the manufacturer’s package insert instructions.^15^ The VITROS CoV2T test is an antigen sandwich immunoassay designed to qualitatively detect antibody to the S1 subunit of the SARS-CoV-2 spike glycoprotein, including IgA, IgM and IgG. The assay involves a two-stage reaction, in which SARS-CoV-2 antibodies present in the sample initially bind with SARS-CoV-2 antigen coated on the test well. Any unbound sample is removed by washing. Subsequently, a horseradish peroxidase (HRP)-labeled recombinant SARS-CoV-2 antigen is added. The HRP-labeled conjugate binds specifically to anti-SARS-CoV-2 already captured on the test well. Unbound conjugate is removed by a subsequent wash step. The bound HRP conjugate is then measured by a luminescent reaction. A reagent containing luminogenic substrates and an electron transfer agent is added to the test wells. The HRP in the bound conjugate catalyzes the oxidation of the luminol derivative, producing light. The electron transfer agent (a substituted acetanilide) increases the level of light produced and prolongs its emission. The light signals are read by the VITROS system. The amount of HRP conjugate bound is indicative of the amount of anti-SARS-CoV-2 antibody present. Results of the test are reported as either Reactive (S/CO ≥ 1.0) or Nonreactive (S/CO < 1.0), although the S/CO can also be obtained from the system.

### Neutralization Testing

Study samples were tested with the SARS-CoV-2 Reporter Viral Particle Neutralization (RVPN) assay at VRI.^16^ Upon receipt serum samples were plated in a 96-well plate and heat-inactivated (HI) for 30 min at 56°C. Reporter viral particles (RVPs) represent a safe and rapid way of quantitatively measuring neutralization, by using SARS-CoV-2 Spike glycoprotein pseudotyped onto a rhabdovirus reporter virus. Neutralization was performed using vesicular stomatitis virus (VSV)-based pseudovirions (PV) as previously described.^16^ Briefly, HEK293T cells were transfected with human ACE2 and TMPRSS2 by TransIT-2020. 24 hours later cells were plated into black 96-well tissue culture treated plates. Four-fold dilutions of heat inactivated serum were performed and VSV-firefly luciferase pseudovirus harboring SARS-CoV-2 S (adjusted to result in 10,000-20,000 RLU in target cells), was mixed with the respective serum dilution, no serum controls and a positive and negative serum control. The mixture was incubated for one hour at 37°C and then used to infect the transfected cells in black plates. After 24 hours at 37°C, supernatant was removed, cells lysed and luciferase activity was measured as per manufacturer’s instructions (Promega, Madison, WI). Fifty percent neutralization titers (NT_50_) were estimated by calculating percent of no serum control and plotting non-linear regression curves constructed in GraphPad Prism version 8.4 (GraphPad Software, San Diego, CA). Donors are considered to lack neutralizing antibodies if their titer was <40.

### Statistical Analysis

CoV2T sensitivity and specificity for selection of CCP at four RVPN NT_50_ threshold nAb titers (1:80, 1:160, 1:320, and 1:640) were determined by dichotomizing the RVPN titer outcomes at the threshold level of interest and preparing receiver operating curve (ROC) analyses using a computer program (NCSS 10 Statistical Software, 2015). NCSS, LLC. Kaysville, Utah, USA). Based on the prevalence of CCP above the RVPN threshold titers, positive and negative predictive values (PPV and NPV, respectively) were calculated. Two-stage CCP release strategies were examined at several CoV2T acceptance values by determining the proportion of CCP within a S/CO range for reflex RVPN that exceed the selected nAb threshold (SAS 9.4, SAS Institute, Cary, NC). A non-parametric Spearman rank correlation coefficient was performed between CoV2T and RVPN (SAS 9.4, SAS Institute, Cary, NC).

## Results

### Donor Demographics

753 CCP donors were tested from three blood centers across the United States (Table 1). Median donor age was 47 years (range 15-84 years) and roughly equally distributed between males and females (48.8% female). ABO blood type frequencies were relatively consistent with previously published prevalences in the US population: group O 39.5%, group A 40.3%, group B 13.4%, group AB 6.8%.^17^ The CCP donors were well represented across the United States hailing from 39 of the 48 continental states (Figure 1). Florida accounted for the entire CCP donor population from one blood center with the other two covering 38 states. Colorado and California had the second two highest study donor populations (16.9% and 8.9% respectively. Several states (i.e. Rhode Island, Wyoming, Alabama, New Mexico) had only one donor representing 0.1% of the overall study population.

**Table 1.** Convalescent Plasma Donor Demographics and Summary Plasma Antibody Test Results

|  | **ARC**<br>**(N=251)** | **Vitalant**<br>**(N=249)** | **OneBlood**<br>**(N=253)** | **Summary**<br>**(N=753)** |
| --- | --- | --- | --- | --- |
| **Age (years) at time of Donation**<br>**(median, range)** | 43 (18-77) | 50 (17-77) | 48 (15-84) | 47 (15-84) |
| **Sex (% F, M)** | 52.4, 47.6 | 49.2, 50.8 | 44.7, 55.3 | 48.8, 51.2 |
| **ABO (%)** | O: 46.4<br>A: 36.8<br>B: 12.4<br>AB: 4.4 | O = 34.4<br>A = 40.2<br>B = 17.3<br>AB = 8.0 | O = 39.9<br>A = 41.5<br>B = 10.7<br>AB = 7.9 | O = 39.5<br>A = 40.3<br>B = 13.4<br>AB = 6.8 |
| **VITROS % reactive (S/Co>1.0)** | 89.2% | 98.0% | 93.3% | 93.5% |
| **VITROS Median (IQR)** | 81.8 (17.6-200) | 82.2 (23.5-163) | 59.7 (18.1-117) | 71.2 (19.9-156) |
| **RVPN Median (IQR)** | 376 (127-937) | 732 (240-1941) | 604 (225-2076) | 528 (182-1627) |
| **RVPN NT50>1:80** | 81.7% | 90.0% | 87.4% | 86.3% |
| **RVPN NT50>1:160** | 67.3% | 81.5% | 80.2% | 76.4% |
| **RVPN NT50>1:320** | 53.8% | 66.7% | 64.4% | 61.6% |
| **RVPN NT50>1:640** | 33.9% | 52.6% | 48.2% | 44.9% |

**Figure 1.**
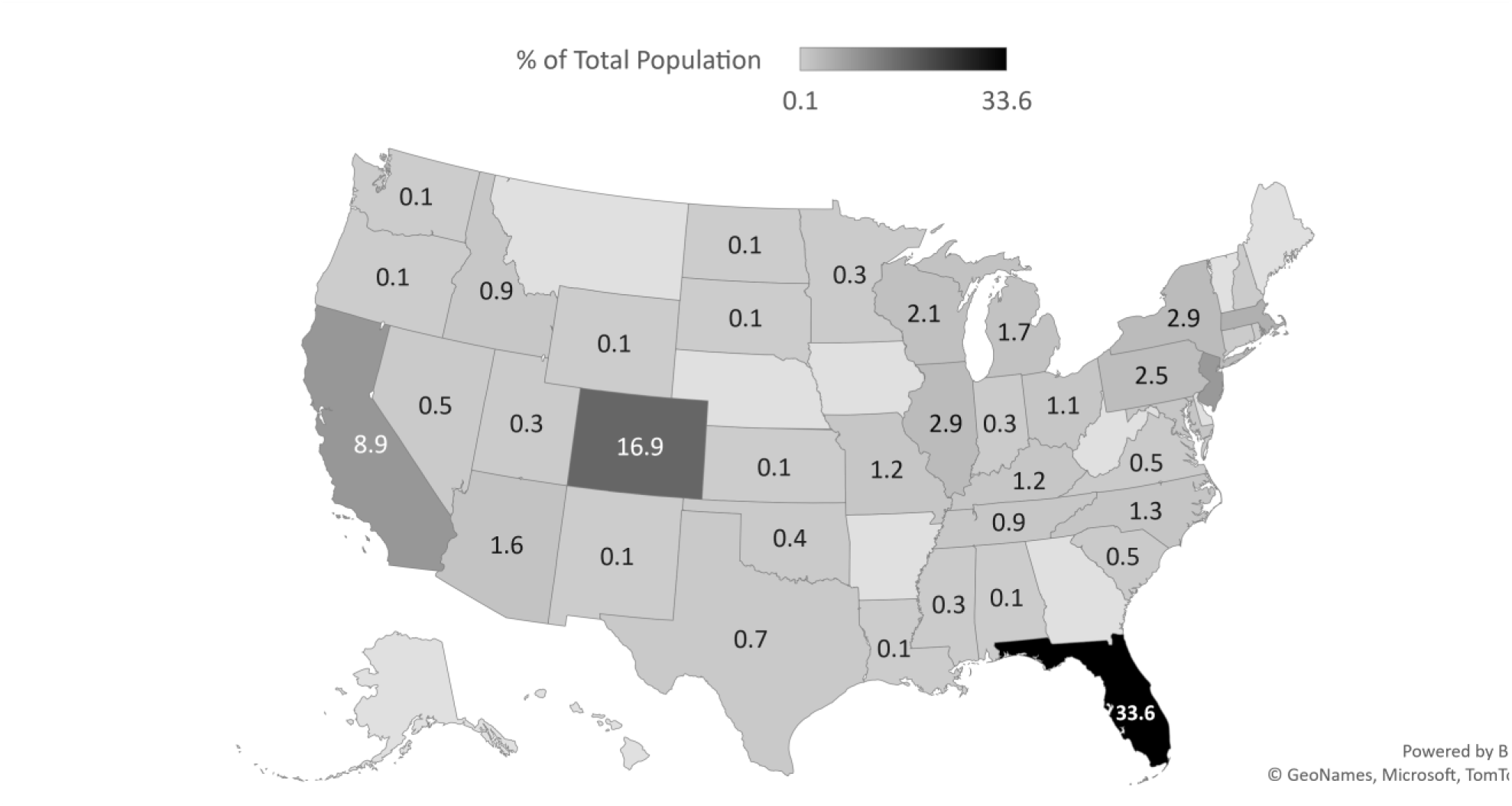
Geographic distribution of CCP donors. 59.4% of CCP donors in the study were from three states: Florida, Colorado, and California.

The ARC samples had a slightly lower percent reactive on the VITROS CoV2T, and a lower median nAb titer in the RVPN. This may have been a consequence of the difference in collection catchments; and would be expected to change as the pandemic peak progresses around the country. This is not relevant for evaluation of the CoV2T predictive performance.

### Population CoV2T and RVPN Testing

The 753 CCP donors had a median CoV2T S/CO of 71.2 (IQR 19.9-156) and a median NT_50_ of 528 (IQR 182-1627) (Table 1). 94% were reactive for CoV2T and had a neutralizing titer >40 with 6% “sero-silent” with nonreactive S/CO and absence of a neutralizing titer. Figure 2 shows the distribution of study population based on CoV2T S/CO results against RVPN titers. The distribution is not normal and suggests a direct linear relationship between CoV2T and RVPN consistent with other studies.^18^ A Spearman rank correlation coefficient showed low correlation between CoV2T and RVPN (r = 0.34217).

**Figure 2.**
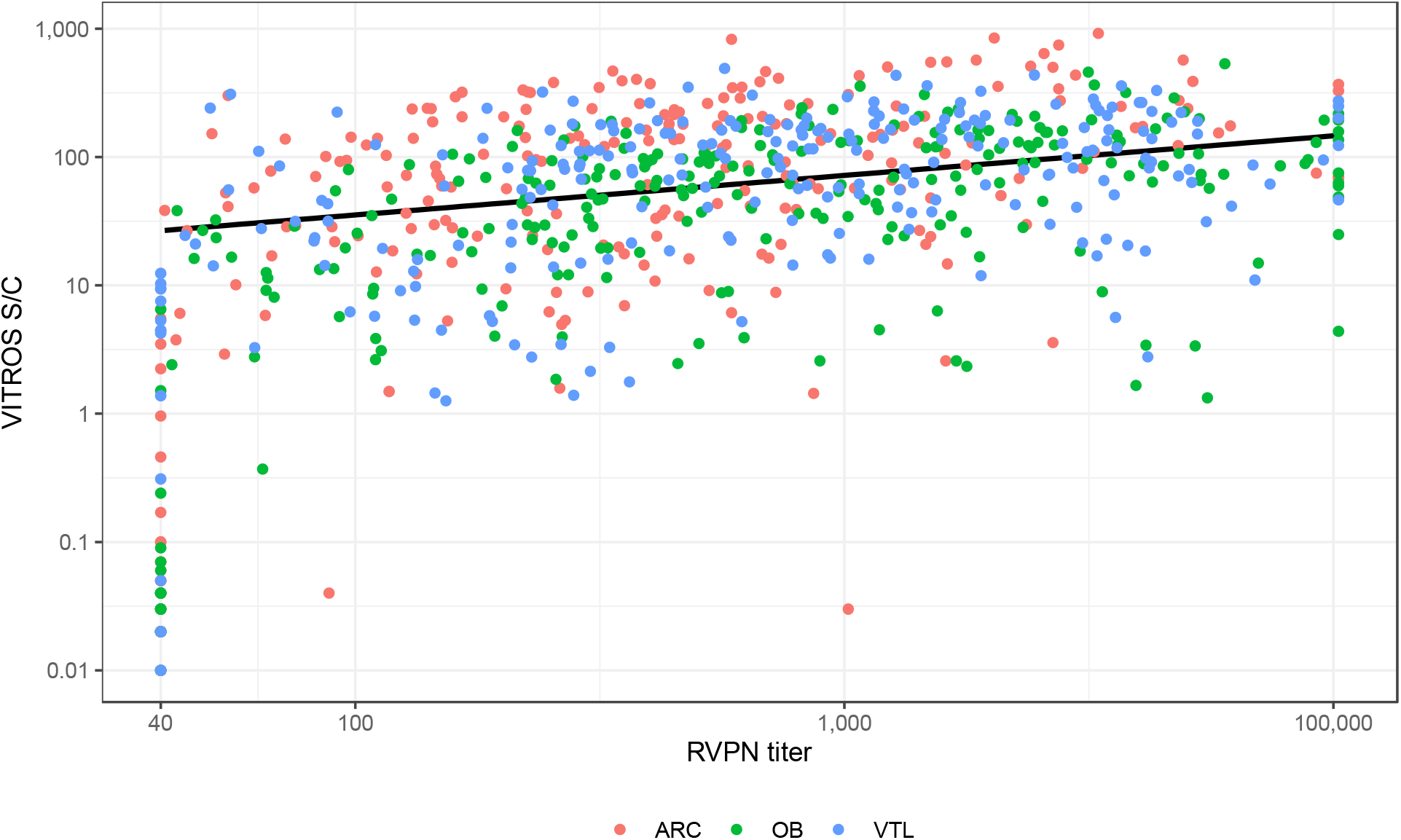
Distribution of CoV2T S/Co versus RVPN NT_50_. The distribution is not normal but suggests a direct linear correlation between the two tests. Truncated RVPN results beyond the assay response range are evident at 1:40 and 1:100,000. Slope and R^2^ of the regression line excluding the truncated results is y = 2.10 + 0.39x with an R^2^ = 0.14.

Table 2 summarizes proportions of reactive CCP donations at S/CO thresholds of 1, 10, 20, 30, and 60 that yields RVPN NT_50_ titers meeting proposed CCP minimum potency levels. As the S/CO threshold approaches 60 the proportion of selected donors meeting candidate NT_50_ titers increases – with 98% of donors with S/CO ≥60 had NT_50_ ≥1:80 and 79% ≥1:320. However, at an S/CO threshold of 60, 45% of donors do not meet the 60 S/CO criterion (Table 2), many of whom have high neutralization titers.

**Table 2.** NT_50_ Titers at Varied CoV2T Thresholds

| **Threshold**<br>**(S/Co)** | **Number (%) Reactive**<br>**CCP Donors ≥Threshold** | **Number (%) of CCP Donors Above NT<sub>50</sub>**<br>**Threshold Titer** |  |  |  |
| --- | --- | --- | --- | --- | --- |
|  |  | **≥1:80** | **≥1:160** | **≥1:320** | **≥ 1:640** |
| **≥1** | 704 (94%) | 648 (92) | 574 (82) | 463 (66) | 337 (48) |
| **≥10** | 619 (82%) | 585 (94) | 527 (85) | 435 (70) | 320 (52) |
| **≥20** | 564 (75%) | 539 (96) | 494 (88) | 411 (73) | 306 (55) |
| **≥30** | 508 (67%) | 492 (97) | 456 (90) | 387 (76) | 287 (56) |
| **≥60** | 414 (55%) | 406 (98) | 383 (93) | 327 (79) | 246 (59) |

The CoV2T ROCs are shown in Figure 3 for NT_50_ thresholds of 1:80, 1:160, 1:320 and 1:640. As the threshold nAb titer criterion increases, the performance of the CoV2T assay for selecting CCP above the threshold deteriorates (AUC 0.88, 0.82, 0.76, 0.71). Potential CoV2T S/CO cutoffs are superimposed on the ROC illustrating how increasing the S/CO criterion results in a tradeoff effect of decreasing sensitivity (probability of a true positive selection) while decreasing the risk of a false positive selection. This is also shown in Table 3 with various combinations of CoV2T cutoff S/CO and desired CCP NT_50_. Based on the prevalence of NT_50_ greater than threshold values, the PPV and NPV for the CoV2T assay illustrate the tradeoff associated with increasing the NT_50_ requirement.

**Table 3.** Performance of CoV2T for Prediction of Neutralizing Antibody Titers – PPV & NPV (positive and negative predictive values) are shown for the prevalence of CCP with NT_50_> the indicated thresholds.

| **NT50**<br>**Threshold** | **PREVALENCE**<br>**≥Threshold** | **S/CO**<br>**Criteria** | **Sensitivity** | **Specificity** | **PPV** | **NPV** |
| --- | --- | --- | --- | --- | --- | --- |
| 80 | 0.86 | 1 | 0.997 | 0.447 | 0.919 | 0.958 |
| 160 | 0.76 | 1 | 0.998 | 0.264 | 0.814 | 0.979 |
| 320 | 0.62 | 1 | 0.998 | 0.163 | 0.657 | 0.979 |
| 640 | 0.45 | 1 | 0.997 | 0.113 | 0.478 | 0.979 |
| 80 | 0.86 | 10 | 0.900 | 0.670 | 0.945 | 0.515 |
| 160 | 0.76 | 10 | 0.917 | 0.483 | 0.851 | 0.642 |
| 320 | 0.62 | 10 | 0.938 | 0.363 | 0.703 | 0.784 |
| 640 | 0.45 | 10 | 0.947 | 0.280 | 0.517 | 0.866 |
| 80 | 0.86 | 20 | 0.832 | 0.757 | 0.956 | 0.417 |
| 160 | 0.76 | 20 | 0.863 | 0.607 | 0.876 | 0.578 |
| 320 | 0.62 | 20 | 0.888 | 0.467 | 0.728 | 0.722 |
| 640 | 0.45 | 20 | 0.905 | 0.373 | 0.541 | 0.829 |
| 80 | 0.86 | 30 | 0.758 | 0.845 | 0.969 | 0.357 |
| 160 | 0.76 | 30 | 0.795 | 0.708 | 0.898 | 0.516 |
| 320 | 0.62 | 30 | 0.836 | 0.581 | 0.762 | 0.689 |
| 640 | 0.45 | 30 | 0.852 | 0.467 | 0.566 | 0.795 |
| 80 | 0.86 | 60 | 0.625 | 0.922 | 0.981 | 0.280 |
| 160 | 0.76 | 60 | 0.666 | 0.826 | 0.925 | 0.434 |
| 320 | 0.62 | 60 | 0.705 | 0.699 | 0.790 | 0.596 |
| 640 | 0.45 | 60 | 0.728 | 0.595 | 0.594 | 0.729 |

**Figure 3.**
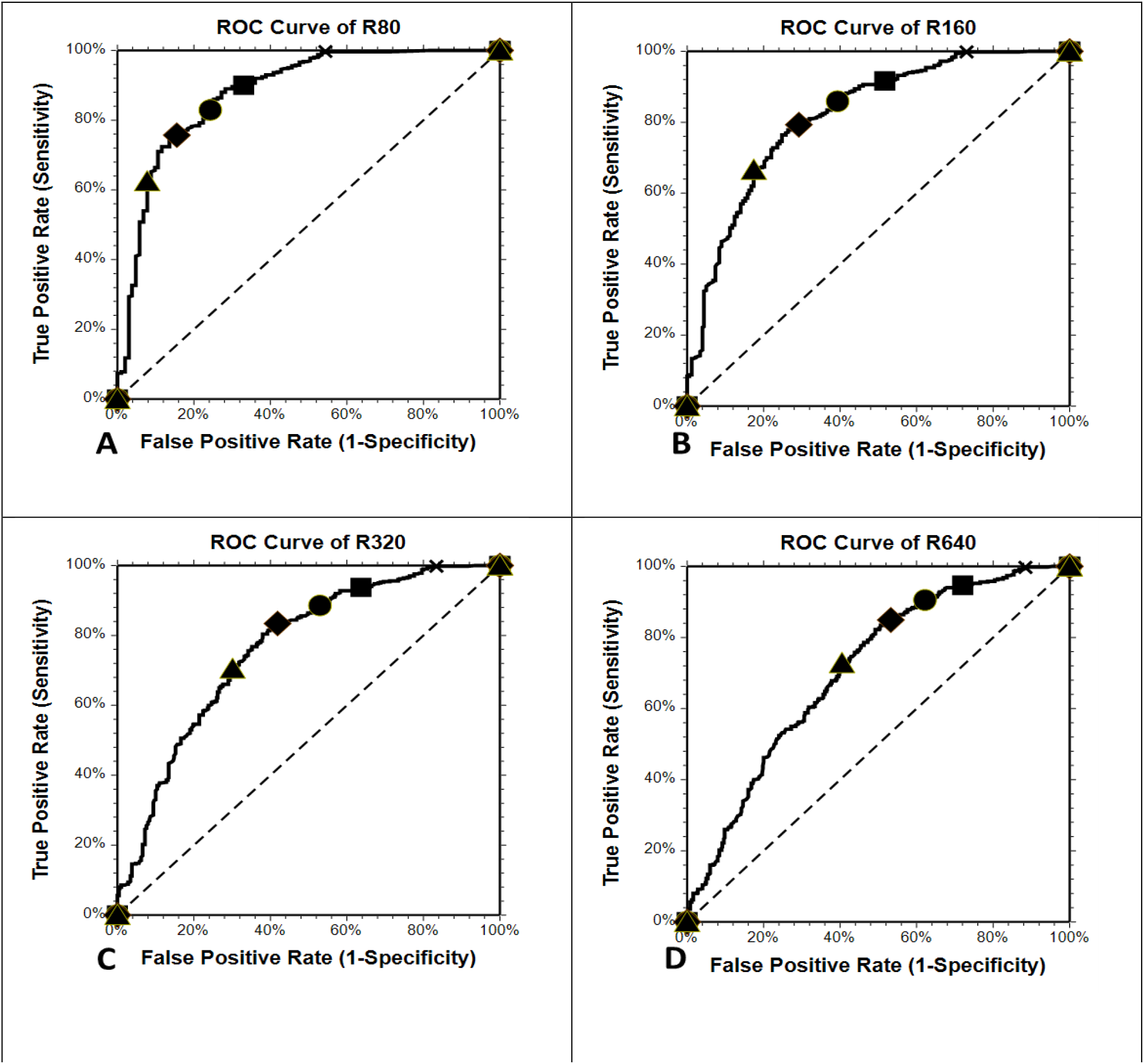
ROC Curves for Varied nAb Titer Thresholds. As the nAb titer increases the ability of the S/Co to predict the nAb titer decreases. ▲:S/CO=60 ♦:S/CO=30 ●:S/CO=20 ■:S/CO=10 X:S/CO=1.

We examined the effect of a two-stage selection process for CCP donations (Figure 4): Stage 1 is a CCP rejection with S/CO<1.0 and immediate acceptance for S/CO>acceptance criterion; Stage 2 is reflexed secondary screening for 1.0<S/CO<acceptance criterion illustrated here as a RVPN assay. The anticipated outcome of several such threshold and acceptance criteria are shown in Table 4. For example, with an S/CO acceptance criteria of 30, 6.5% are immediately rejected with S/CO<1.0, 67.5% are immediately accepted as CCP, and 26% are reflexed to secondary screening. With a NT_50_ threshold of 1:160, 89.8% with S/CO>30 are anticipated to be at or above 1:160 and 60.2% of the reflexed tested units would be releasable with RVPN NT_50_>1:160. This results in 91.7% of the final released inventory expected to have NT_50_>1:160. Approximately 11% of the all tested CCP would be rejected and not released as CCP.

**Table 4.** CCP unit reflex testing using Two-stage method: CoV2T and RVPN – A) STAGE 1 – Initial Screen: Selects units that are immediately partitioned to FP24 or CCP based on CoV2T S/CO threshold. N (% of total) B) STAGE 2 – Retesting: Reflexes unit in the intermediate zone to RVPN testing for partitioning based on NT_50_ requirements. FP24: NT_50_ <titer; CCP: NT_50_ ≥titer; N (% of retested)

| A) STAGE 1 – Initial Screen with CoV2T |  |  |  |  |  |  |  |  |
| --- | --- | --- | --- | --- | --- | --- | --- | --- |
| S/CO Threshold | 10 |  | 20 |  | 30 |  | 60 |  |
| Qualified as CCP<br>S/CO ≥ threshold | 619 (82.2%) |  | 564 (74.9%) |  | 508 (67.5%) |  | 414 (55%) |  |
| Qualified as PF24<br>S/CO < threshold | 49 (6.5%) |  | 49 (6.5%) |  | 49 (6.5%) |  | 49 (6.5%) |  |
| Reflex Testing<br>1.0≤S/CO<threshold | 85 (11%) |  | 140 (19%) |  | 196 (26%) |  | 290 (39%) |  |
| B) STAGE 2 – Reflex Testing with RVPN |  |  |  |  |  |  |  |  |
| RVPN Titer (NT<sub>50</sub>) | FP24 | CCP | FP24 | CCP | FP24 | CCP | FP24 | CCP |
| ≥1:80 | 22 (25.9) | 63 (74.1) | 31 (22.1) | 109 (77.9) | 40 (20.4) | 156 (79.6) | 48 (16.6) | 242 (83.5) |
| ≥1:160 | 38 (44.7) | 47 (55.3) | 60 (42.9) | 80 (57.1) | 78 (39.8) | 118 (60.2) | 99 (34.1) | 191 (65.9) |
| ≥1:320 | 57 (67.1) | 28 (32.9) | 88 (62.9) | 52 (37.1) | 120 (61.2) | 76 (38.8) | 154 (53.1) | 136 (46.9) |
| ≥1:640 | 68 (80) | 17 (20.0) | 109 (77.9) | 31 (22.1) | 146 (74.5) | 50 (25.5) | 199 (68.6) | 91 (31.4) |

**Figure 4.**
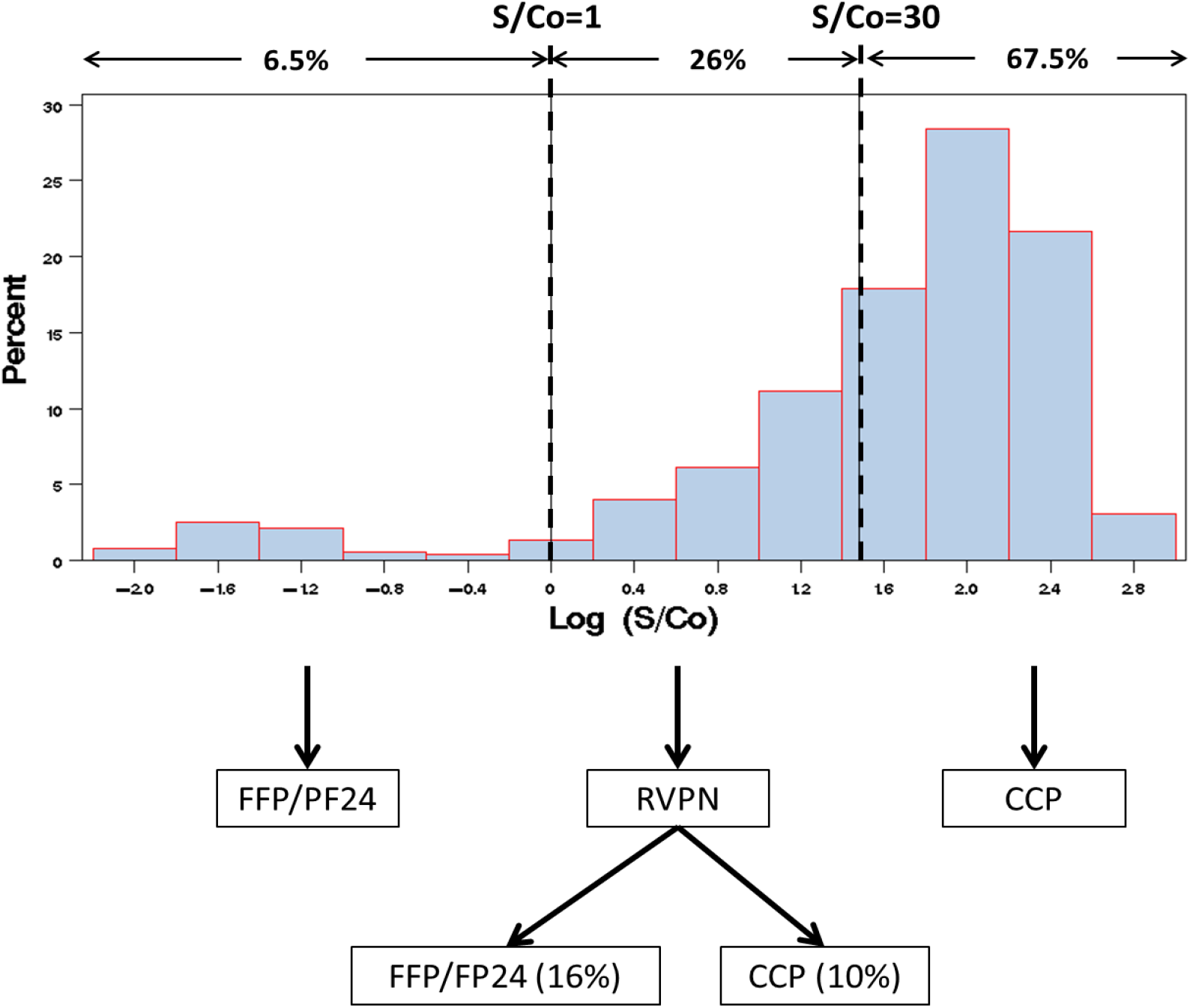
Reflex Testing Example - S/CO=30 as CoV2T acceptance criteria and target NT_50_>320. STAGE 1: Release as CCP with S/CO≥30 and release as FFP/FP24 with S/CO<1.0. STAGE 2 (reflex testing): 1.0≤S/CO<30 – 26% of units are tested with RVPN; 10% of total (38% of retested) units meet the target RVPN threshold.

## Discussion

COVID-19 has presented new and daunting challenges for our society worldwide. The last six months have seen incredible strides in public health and prevention as well as understanding of disease pathogenesis and formulation of effective treatment protocols. Viral specific therapy is still in development setting the stage for temporary use of an historical treatment: convalescent plasma. Initial studies using CCP have suggested efficacy particularly when used earlier in the disease course though the most efficacious nAb titer and dose has not yet been determined.^6,19^

Although bAbs may protect through multiple effector pathways, the effectiveness of passive immunotherapy is believed to be broadly proportional to the potency of the intervention as determined with nAb titers. As applied to CCP on the large-scale experience in the US, it is not possible to provide timely screening of CCP products due to technological limitations. Therefore, it is imperative to apply high throughput bAb assays as indicators of nAb levels. We have demonstrated that the CoV2T test system, while not a perfect predictor of nAb titers, is capable of identifying donors with no Ab response. The test system also presents a S/CO readout with a large dynamic range. This output range can be exploited to provide reasonable PPV as a screening tool. For example, a S/CO criterion of 30 applied to a previously qualified CCP donor population is expected to result in 90% of CCP with NT_50_ ≥1:160. We further illustrated that reflex testing of CCPP in the intermediate zone of S/CO 1-30 could provide additional units at or above the desired NT_50_. Such an approach will permit high throughput screening for timely provision of CCP.

The selection of targeted nAb titers for clinical use will significantly affect the yield and availability of CCP transfusion. Due to inherent limitations of large volume testing for neutralization titers, we present a potential reflex testing algorithm using initial high-throughput CoV2T testing. The S/CO thresholds selected for routine product release will need to balance the risk of releasing products without the minimum target nAb titer. Organizations may consider this testing scheme or develop others to effectively select for CCP. Another approach that could be considered is pooling of CCP units from different donors. This would narrow the distribution of nAb titers in the CPP and take advantage of regression to the mean. In other words, the lower titer units would be balanced by the higher titer units and result in higher proportions of pooled CCP above any desired nAb titer. Regardless, any algorithm would need to effectively evaluate testing performance characteristics including sensitivity and specificity as well as PPV and NPV while correlating with clinical outcomes.

Our study did have several limitations. The SARS-CoV-2 RVPN detects the presence of neutralizing antibody directed against the virus, but there is still a potential that cross-reactivity with other coronavirus species may occur. In addition, though represented in 39 different states, the study population was not entirely representative of the potential CCP donor pool within the

United States which as of August 25, 2020 exceeds 124, 036.^1,17^ Despite being the largest published report of CCP donors to date, our population was still relatively small and collected in the early days of the pandemic. Our cross-sectional analysis showed 94% seropositivity to both the CoV2T and the RVPN with 6% sero-silent consistent with a recent serosurvey of recovered COVID-19 patients.^20^ However, our testing only examined humoral response to the immune dominate S-protein. Additional studies are necessary to examine humoral response to additional viral antigens and correlate with days from COVID-19 symptom onset and severity of illness.

Ab reactivity on the CoV2T testing system appears to be quite stable over at least 90 days (unpublished), however, other assay platforms targeting Ig-only suggest waning for the IgG response, and possibly, nAb as well.^21,22^ Waning of nAb over time when the CoV2T test presents a stable signal strength would reduce the utility of this system as a predictor of nAb and selection of CCP for transfusion. Though not directly applicable to recent FDA EUA guidance regarding CCP unit test, the information obtained from correlating these two testing platforms is helpful in directing specific reflex testing for donors not meeting initial testing criteria.

## Data Availability

n/a

## References

1. Coronavirus Resource Center, John’s Hopkins University and Medical Center. [cited 2020 Aug 25]. Available from: https://coronavirus.jhu.edu/map.html.

2. Investigational COVID-19 Convalescent Plasma: Guide for Industry, the Food and Drug Administration. [cited 2020 July 27]. Available from: https://www.fda.gov/media/136798/download.

3. Shen C, Wang Z, Zhao F, et al. Treatment of 5 Critically Ill Patients With COVID-19 With Convalescent Plasma. JAMA 2020;323(16):158209. doi: 10.1001/jama.2020.4783.

4. Duan K, Liu B, Cesheng L, et al. Effectiveness of convalescent plasma therapy in severe COVID-19 patients. Proc Natl Acad Sci USA 2020;117(17):9490–9496. doi: 10.1073/pnas.2004168117.

5. Salazar E, Perez KK, Ashraf M, et al. Treatment of COVID-19 Patients with Convalescent Plasma. Am J Pathol. 2020;190(8): 1680–1690. doi: 10.1016/j.ajpath.2020.05.014.

6. Joyner MJ, Senefeld JW, Klassen SA, et al. Effect of Convalescent Plasma on Mortality among Hospitalized Patients with COVID-19: Initial Three-Month Experience. medRxiv preprint published 2020 Aug 12. doi:10.1101/2020.08.12.20169359.

7. Joyner MJ, Wright RS, Fairweather D, et al. Early safety indicators of COVID-19 convalescent plasma in 5000 patients. J Clin Invest 2020;140200. doi:10.1172/JCI140200. Online ahead of print.

8. Li L, Zhang W, Hu Y, et al. Effect of Convalescent Plasma Therapy on Time to Clinical Improvement in Patients with Severe and Life-threatening COVID-19: A Randomized Clinical Trial. JAMA 2020; 4;324(5):519. doi:10.1001/jama.2020.10044

9. Hung IF, To KK, Lee CK, et al. Convalescent plasma treatment reduced mortality in patients with severe pandemic influenza A (H1N1) 2009 virus infection. Clin Infect Dis 2011;52(4):447–456.

10. Ko JH, Seok H, Cho SY, et al. Challenges of convalescent plasma infusion therapy in Middle East respiratory coronavirus infection: a single centre experience. Antivir Ther 2018;23(7):617–622.

11. VanBlargan LA, Goo L, Pierson TC. Deconstructing the Antiviral Neutralizing-Antibody Response: Implications for Vaccine Development and Immunity. Microbiol Mol Biol Rev 2016;80(4):989–1010.

12. Klasse PJ, Moore JP. Antibodies to SARS-CoV-2 and their potential for therapeutic passive immunization. Elife 2020;23. doi:10.7554/eLife.57877.

13. Recommendations for Investigational COVID-19 Convalescent Plasma, the Food and Drug Administration. Cited on 2020 Aug 24. Available from: https://www.fda.gov/media/141480/download.

14. Code of Federal Regulations. Title 21, CFR Parts 600 to 799. Washington, DC: US Government Printing Office, 2019 (revised annually).

15. Ortho Clinical Diagnostics. VITROS Immunodiagnostic Products Anti-SARS-CoV-2 Total Reagent Pack and VITROS Immunodiagnostic Products Anti-SARS-CoV-2 Total Calibrator Instructions for Use. Version 3.0. Ortho Clinical Diagnostics, Inc. Rochester, New York. April 14, 2020.

16. Ng D, Goldgof G, Shy B, et al. SARS-CoV-2 seroprevalence and neutralizing activity in donor and patient blood from San Francisco Bay Area. medRxiv preprint published online May 2020. doi: 10.1101/2020.05.19.20107482.

17. AABB, Blood FAQ. [cited 2020 Aug 25]. Available from: http://www.aabb.org/tm/Pages/bloodfaq.aspx.

18. Luchsinger LL, Ransegnola B, Jin D, et al. Serological analysis of New York City COVID-19 Convalescent Plasma Donors. medRxiv preprint published online June 2020. doi: 10.1101/2020.06.08.20124792.

19. Piechotta V, Chai KL, Valk SJ, et al. Convalescent plasma or hyperimmune immunoglobulin for people with COVID-19: a living systematic review. Cochrane Database Syst Rev. Published online 10 July 2020. doi:10.1002/14651858.CD013600.pub2.

20. Petersen LR, Sami S, Vuong N, et al. IgG antibodies to SARS-CoV-2 in a large cohort of previously infected persons. JAMA Internal Medicine 2020, In review.

21. Wajnberg A, Amanal F, Firpo A, et al. SARS-CoV-2 infection induces robust, neutralizing antibody responses that are 1 stable for at least three months. medRxiv preprint published online July 2020. doi: https://doi.org/10.1101/2020.07.14.20151126

22. Seow J, Graham C, Merrick B, et al. Longitudinal evaluation and decline of antibody responses in SARS-CoV-2 infection. medRxiv preprint published online July 2020. doi: https://doi.org/10.1101/2020.07.09.20148429.

